# Empiric Dietary Inflammatory Potential Score, Inflammatory Biomarkers, and Risk of Atrial Fibrillation: The Atherosclerosis Risk in Communities Study

**DOI:** 10.64898/2026.02.23.26346939

**Authors:** Mohamed Mostafa, Matthew J. Singleton, Ghanshyam P. Shantha, Prashant D. Bhave, Joseph Yeboah, Elsayed Z. Soliman, Takeki Suzuki

## Abstract

**Background:** Inflammation plays a key role in atrial fibrillation (AF) pathogenesis. The empirical dietary inflammatory potential (EDIP) score predicts circulating inflammatory biomarkers and adverse cardiac outcomes, but its association with incident AF is unclear. This study aimed to examine the relationship between EDIP score and AF risk.

**Methods:** Participants from the Atherosclerosis Risk in Communities (ARIC) free of baseline AF who completed a validated food frequency questionnaire were included. Correlation of EDIP with inflammatory biomarkers (factor VIII, fibrinogen, von Willebrand factor, and C-reactive protein) was examined at baseline. Incident AF was ascertained using electrocardiograms, hospital records, and death certificates. Cox proportional hazards models estimated hazard ratios of AF across EDIP quantiles and per SD increase, adjusting for sociodemographic and cardiovascular risk factors.

**Results:** Among 8,277 participants (54.1 years old, 51.3% women, 80% white), higher EDIP score correlated with circulating inflammatory biomarkers at baseline. Over a median 24.2 years of follow-up, 1,453 had incident AF (incident rate 8.6 per 1,000 person-years). Compared with the most anti-inflammatory diet (EDIP Q1), the most pro-inflammatory diet (EDIP Q5) was associated with increased AF risk (HR 1.21; 95% CI 1.03–1.43). Sex-stratified analyses showed a stronger association in men (HR 1.43; 95% CI 1.14–1.79), while no significant association was observed in women.

**Conclusions:** Pro-inflammatory dietary patterns are independently associated with higher AF risk in a middle-aged cohort. These findings would support incorporating dietary inflammatory load into AF risk stratification.

**Clinical Perspective What Is New?:**

- Higher Empirical Dietary Inflammatory Potential (EDIP) scores, indicating a more pro inflammatory diet, were associated with an increased long-term risk of atrial fibrillation (AF) in a large, biracial, community-based cohort followed for over two decades.
- Sex stratified analyses revealed a significant sex difference: higher EDIP scores were consistently associated with increased AF risk in men, whereas no significant association was observed in women, suggesting sex-specific susceptibility to EDIP.
- Obesity modified the association between EDIP and AF, with the strongest risk observed among individuals with BMI ≥30, while an inverse or attenuated association was seen among normal weight participants.

**What Are the Clinical Implications?:**

- Dietary inflammatory load may serve as a meaningful and modifiable upstream AF risk factor, complementing conventional cardiovascular risk assessment, particularly in men and individuals with obesity.
- Incorporating dietary pattern assessment into routine AF risk stratification may help identify individuals who could benefit most from targeted lifestyle interventions.
- Public health and clinical prevention strategies promoting anti-inflammatory dietary patterns (e.g., increased intake of fruits, vegetables, and whole grains; reduced intake of processed meats and refined carbohydrates) could meaningfully reduce AF incidence.
- Recognition of sex specific differences in AF pathways reinforces the need for personalized preventive strategies, as diet inflammation mechanisms appear to influence AF development more prominently in men.

## INTRODUCTION

Atrial fibrillation (AF) is the most common sustained cardiac arrhythmia and a major contributor to cardiovascular morbidity, mortality, and impaired quality of life.^1^ Its prevalence increases sharply with age, affecting nearly 5% of adults over 65 years, and is projected to rise substantially with population aging.^2,3^ with sex-related heterogeneity in its risk factors, clinical course, and outcomes.^4–6^ Although men have a higher lifetime risk and earlier onset of AF, women tend to develop AF at older ages, experience greater symptom burden, and have higher risks of stroke, heart failure, and mortality once AF occurs.^7,8^ Women also demonstrate different patterns of AF risk factors, with stronger associations of hypertension, valvular disease, and obesity. ^6,9,10^

Contemporary guidelines emphasize AF risk factor modification and lifestyle interventions, particularly weight loss, as key components of AF management.^11–13^ However, the contribution of lifestyle factors beyond weight, especially dietary patterns, to AF risk remains incompletely understood. Beyond traditional metabolic mechanisms, habitual dietary patterns play a particularly important role in shaping systemic inflammatory burden over time.^14–18^ The Empirical Dietary Inflammatory Potential (EDIP) score provides a validated measure of the inflammatory properties of habitual diets, capturing their long-term impact on circulating inflammatory biomarkers and cardiovascular risk.^19^ Higher EDIP scores, indicative of pro-inflammatory dietary patterns, have been independently linked to adverse cardiovascular outcomes and increased mortality.^20–22^

Despite these insights, the relationship between EDIP and incident AF remains unclear. We hypothesize that higher EDIP scores, reflecting more pro-inflammatory dietary patterns, are associated with an increased risk of AF beyond traditional cardiovascular risk factors in the Atherosclerosis Risk in Communities (ARIC) cohort, a large, community-based biracial population.

## METHODS

### Study Population

The design and conduct of the ARIC study has been reported previously^.23^ Briefly, ARIC is a prospective cohort of 15,792 persons, aged 45 to 64 years at the time of enrollment (1987 – 1989), to understand in more detail the development of cardiovascular diseases and their risk factors in the general population. It recruited from four United States communities: Forsyth County, North Carolina; Jackson, Mississippi; the northwestern suburbs of Minneapolis, Minnesota; and Washington County, Maryland. The institutional review boards from each of the ARIC study sites, as well as the Coordinating Center, approved the study protocols. This analysis was conducted using de-identified data in accordance with the Research Materials Distribution Agreement (RMDA) between the authors and the NHLBI BioLINCC repository. This manuscript was prepared using a limited access dataset obtained from BioLINCC. All participants provided written informed consent at the time of data collection. We excluded those with AF on electrocardiogram (ECG) at the baseline visit, those who did not have a baseline ECG, and those with incomplete dietary self-report data, leaving a total of 8,277 participants.

### Exposure Variable

At the baseline interview, participants’ dietary patterns were assessed using a modified version of a well-validated food frequency questionnaire (FFQ).^24^ This FFQ included 66 categories of food and drink and was administered by trained interviewers. Participants were asked to quantify their average consumption of each category of food and drink over the preceding year, with nine categories of frequency, ranging from “almost never” to “more than six times per day.” To allow for modeling of food type consumption as a continuous variable, the categories of consumption were transformed into servings per day according to the following weights: “almost never” = 0, “1 – 3 per month” = 0.066, “1 per week” = 0.14, “2 – 4 per week” = 0.43, “5 – 6 per week” = 0.79, “1 per day” = 1.0, “2 – 3 per day” = 2.5, “4 – 6 per day” = 5.0, and “more than 6 per day” = 7.0.^25^ The EDIP score was calculated based on the inflammatory potential of 18 food groups derived from this questionnaire. Each food group was expressed as servings per day and multiplied by its corresponding empirically derived coefficient, reflecting its association with circulating inflammatory markers. The weighted values of these 18 food groups were summed to generate an overall EDIP score. Additional details regarding score derivation and validation have been published previously.^19^ Similar to prior studies, all scores were divided by 1000 to create simpler values for interpretation and reporting. ^21,22,26^ EDIP score was categorized into quantiles with participants in the fifth quartile having the highest EDIP score indicating a higher pro-inflammatory dietary burden, while those in the first quantile having the lowest score indicating an anti-inflammatory dietary pattern.

### Covariates

Baseline covariates were obtained at the initial ARIC examination (1987 – 1989). Age, sex, race/ethnicity, education level, alcohol intake, cigarette smoking status and medication usage were self-reported at the baseline visit. Body mass index (BMI) was calculated as weight in kilograms divided by the square of height in meters. Hypertension was defined as a systolic blood pressure above 140 mmHg, diastolic blood pressure above 90 mmHg, or the use of antihypertensive medication in the preceding two weeks. Diabetes mellitus was defined as a fasting glucose concentration ≥ 126 mg/dL, non-fasting glucose concentration ≥ 126 mg/dL, self-reported physician diagnosis, or the use of hypoglycemic medication. Prior cardiovascular diseases were defined form self-reported physician diagnosis of heart failure, myocardial infarction or stroke. Levels of total cholesterol (mg/dL) and LDL cholesterol (mg/dL) are collected via phlebotomy during the baseline visit.

Inflammatory biomarkers including fibrinogen, factor VIII (VIII), and von Willebrand factor (vWF) antigen were measured at the ARIC Central Hemostasis Laboratory (University of Texas Medical School, Houston, TX) using standardized ARIC coagulation assays performed on citrate plasma collected in 4.5-mL sodium-citrate tubes during the baseline Visit 1 examination (1987–1989).^27,28^

C-reactive protein (CRP) was measured retrospectively from stored Visit 1 serum aliquots using a high-sensitivity ELISA (Magiwel CRP assay, United Biotech Inc., Mountain View, CA), with a lower detection limit <1 µg/L, a minimum detectable concentration of 0.35 µg/L, and high reproducibility (duplicate r = 0.96). After ARIC had identified the case and cohort samples, technicians retrieved stored samples frozen at −70 °C, and CRP was measured and reported retrospectively from laboratory assays performed years after the baseline visit. ^29^

### Outcome Variable

Our outcome was incident AF. Incident AF was ascertained from a combination of study ECG at ARIC visits 2 through 6, hospital discharge records, and death certificates. Discharge records and death certificates were reviewed and the presence of International Classification of Disease Ninth Edition clinical modification codes 427.31 (AF) or 427.32 (atrial flutter) or I48. in the Tenth Edition in any position in a patient with no history of AF was defined as incident AF. Incident AF events occurring in the same hospitalization as open cardiac surgery were excluded. Prior studies have shown high agreement between AF as ascertained by code and by physician manual chart review.^30^

### Statistical Analysis

Baseline characteristics of the study population stratified by EDIP score were compared using frequency (percentage) for categorical variables and mean ± standard deviation for continuous variables if normally distributed or median and interquartile range if skewed.

Between-group differences were assessed using analysis of variance for continuous variables and chi-squared tests for categorical variables.

To examine the biological relevance of EDIP as a marker of systemic inflammation, we evaluated the cross-sectional correlation between EDIP score and circulating inflammatory biomarkers, including factor VIII, fibrinogen, vWF, and C-reactive protein (CRP). Biomarkers were standardized to z-scores to facilitate comparison across measures. Mean differences in biomarker levels and 95% confidence intervals (CI) were estimated across EDIP quantiles, using the lowest quantile (Q1, most anti-inflammatory diet) as the reference category. Linear trend across EDIP quantiles was assessed by modeling the median EDIP value within each quantile as a continuous term. All models were adjusted for age, sex, race/ethnicity, education and common cardiovascular risk factors including alcohol intake, smoking status, BMI, hypertension, prior CVD (stroke, coronary heart disease, heart failure, myocardial infarction), LDL cholesterol, total cholesterol, antihypertensive medication and intake of lipid lowering medications.

For longitudinal analyses, Non-linear associations between EDIP and risk of AF were explored non-parametrically using restricted cubic splines modeling, with three knots placed at the 10^th^, 50^th^, and 90^th^ percentiles, as recommended by Harrell.^31^ A p-value for non-linearity was computed by testing the null hypothesis that the estimated coefficient of the second spline is zero.^32^ Cox proportional hazards models were used to compare the risk of incident AF as a function of EDIP score, generating hazard ratios (HRs) and 95% CI. The assumption of time-independent proportionality of risks was assessed by examining the Martingale residual plots and by incorporating the natural logarithm of follow-up time as a time-dependent covariate.

For cox proportional hazards models, initial model was unadjusted, with subsequent models adjusted for covariates of clinical significance. Model 1 adjusted for age, sex, race/ethnicity, and education level, while Model 2 adjusting for covariates in Model 1 plus alcohol intake, current smoking, use of lipid lowering meds, antihypertensive medication use, body mass index, LDL cholesterol, total cholesterol, diabetes, hypertension and prior cardiovascular disease (stroke, coronary heart disease, heart failure).

All statistical analyses were performed using The Jamovi project (2025) (Version 2.4.14.0), retrieved from https://www.jamovi.org. Visualization was generated using R version 4.5.1. A two-sided P value of 0.05 was used for hypothesis testing.

## RESULTS

Among 8277 participants (54.1 years old, 51.3% women, 80% white), EDIP score ranged from -1.66 (most anti-inflammatory) to 5.14 (most pro inflammatory) with a mean EDIP score of 0.455. Baseline demographic and clinical characteristics according to EDIP quantiles are presented in **Table 1**. Participants with the highest EDIP score (most pro inflammatory) tended to be males, lower proportion of White participants, have lower educational attainment, higher body mass index (BMI), a higher prevalence of hypertension and diabetes, and greater use of antihypertensive medications.

**Table 1:** Population Characteristics.

| n(%) or<br>(mean ± SD) | Total<br>N=8277 | EDIP Q1<br>N=1665 | EDIP Q2<br>N=1660 | EDIP Q3<br>N=1653 | EDIP Q4<br>N=1650 | EDIP Q5<br>N=1649 | *p*-<br>*value* |
| --- | --- | --- | --- | --- | --- | --- | --- |
| Age | 54.01 (5.75) | 53.78 (5.68) | 54.18 (5.84) | 54.18 (5.64) | 53.93 (5.78) | 54 (5.82) | 0.202 |
| Female | 4247 (51.3) | 869 (52.2) | 856 (51.6) | 879 (53.2) | 856 (51.9) | 787 (47.7) | 0.021 |
| Race: White | 6619 (80.0) | 1403 (84.3) | 1373 (82.7) | 1330 (80.5) | 1314 (79.6) | 1199 (72.7) | <0.001 |
| Education<br>level: |  |  |  |  |  |  | <0.001 |
| Less than<br>high school<br>degree | 1575 (19.0) | 277 (16.6) | 290 (17.5) | 301 (18.2) | 313 (19.0) | 394 (23.9) |  |
| High school,<br>GED, or<br>vocational<br>school | 3507 (42.4) | 725 (43.5) | 689 (41.5) | 688 (41.6) | 714 (43.3) | 691 (41.9) |  |
| College,<br>graduate, or<br>professional<br>school | 3195 (38.6) | 663 (39.8) | 681 (41.0) | 664 (40.2) | 623 (37.8) | 564 (34.2) |  |
| Smoking<br>Status: |  |  |  |  |  |  | 0.001 |
| Current | 1,821 (21.9) | 361 (21) | 332 (20) | 378 (22.9) | 297 (18) | 453 (27.4) |  |
| Former | 3,145 (37.9) | 627 (37.6) | 628 (37.8) | 629 (38) | 630 (38.1) | 631 (38.2) |  |
| Never | 3,311 (40) | 649 (41.6) | 712 (42.8) | 673(40.7) | 677 (41) | 600(36.3) |  |
| Alcohol<br>intake: |  |  |  |  |  |  | 0.172 |
| Current<br>drinker. | 6319 (76.3) | 1248 (75.0) | 1309 (78.9) | 1240 (75.0) | 1259 (76.3) | 1263 (76.6) |  |
| Former<br>drinker | 798 (9.6) | 177 (10.6) | 149 (9.0) | 170 (10.3) | 155 (9.4) | 147 (8.9) |  |
| Never<br>drinker | 1152 (13.9) | 239 (14.4) | 199 (12.0) | 240 (14.5) | 236 (14.3) | 238 (14.4) |  |
| EDIP score | 0.46 (0.43) | -0.09 (0.21) | 0.23 (0.06) | 0.42 (0.05) | 0.46 (0.43) | 0.63 (0.07) | <0.001 |
| BMI | 27.30 (5.08) | 26.93 (4.96) | 27.00 (4.89) | 27.19 (5.01) | 27.56 (5.17) | 27.30<br>(5.08) | <0.001 |
| HTN meds | 2251 (27.2) | 427 (25.6) | 416 (25.1) | 446 (27.0) | 481 (29.2) | 481 (29.2) | 0.039 |
| Lipid<br>lowering | 222 (2.7) | 56 (3.4) | 54 (3.3) | 42 (2.5) | 35 (2.1) | 35 (2.1) | 0.036 |
| Heart failure | 344 (4.2) | 56 (3.4) | 65 (3.9) | 61 (3.7) | 82 (5.0) | 80 (4.9) | 0.075 |
| MI | 318 (3.8) | 55 (3.3) | 58 (3.5) | 70 (4.2) | 61 (3.7) | 318 (3.8) | 0.239 |
| CHD | 376 (4.5) | 66 (4.0) | 74 (4.5) | 79 (4.8) | 74 (4.5) | 83 (5.0) | 0.653 |
| Stroke | 123 (1.5) | 28 (1.7) | 20 (1.2) | 31 (1.9) | 16 (1.0) | 28 (1.7) | 0.422 |
| Hypertension | 2201 (26.6) | 403 (24.2) | 379 (22.8) | 442 (26.7) | 471 (28.5) | 506 (30.7) | <0.001 |
| Diabetes Mellitus | 606 (7.3) | 85 (5.1) | 109 (6.6) | 102 (6.2) | 141 (8.5) | 169 (10.2) | <0.001 |
| Total Cholesterol mg/dL | 215 (42.1) | 216 (42.2) | 214 (41.6) | 217 (40.9) | 215 (42.3) | 215 (43.4) | 0.507 |
| LDL mg/dL | 133.56 (44.19) | 134.65 (44.41) | 132.71 (43.34) | 134.26 (43.61) | 134.21 (44.11) | 131.97 (45.43) | 0.349 |
| Inflammatory Markers: |  |  |  |  |  |  |  |
| CRP mg/L: | 4.41 (9.10) | 4.07 (7.06) | 3.75 (5.92) | 4.52 (10.96) | 4.63 (9.51) | 5.24 (11.22) | 0.040 |
| Factor VIII IU/dL | 128 (36.8) | 125.61 (35.92) | 126.70 (35.50) | 127.72 (34.87) | 128.56 (36.31) | 133.66 (40.86) | <0.001 |
| VWF IU/dL | 115 (46.0) | 115.00 (46.0) | 111.67 (44.47) | 112.61 (42.90) | 114.47 (44.38) | 116.28 (47.06) | <0.001 |
| Fibrinogen mg/dL | 299 (62.9) | 299.00 (62.9) | 297.26 (63.58) | 296.26 (63.34) | 298.14 (61.69) | 301.46 (61.53) | 0.002 |
| EDIP = empirical dietary inflammatory potential. MI: Myocardial Infarction. CHF: coronary heart disease. BMI: body mass index. CRP: C-reactive protein., VWF: von Willebrand factor antigen |  |  |  |  |  |  |  |

### Correlation between EDIP and Inflammatory Markers at Baseline

Participants in the highest quantile of EDIP had higher levels of CRP, factor VIII, VWF and fibrinogen. When examining the correlation between EDIP score and inflammatory markers, higher EDIP scores were strongly and progressively associated with increased levels of VIII, fibrinogen, vWF, and CRP **(Figure 1**). Compared with participants in the lowest EDIP quantile, those in higher quantiles exhibited stepwise increases in standardized levels of factor VIII, fibrinogen, vVF, and CRP. Participants in the highest EDIP quantile (Q5) had the greatest elevations across all biomarkers, with effect sizes exceeding 0.3 standard deviations for factor VIII and vWF. Tests for linear trend across EDIP quantiles were statistically significant for all biomarkers (p for trend < 0.001).

**Figure 1:**
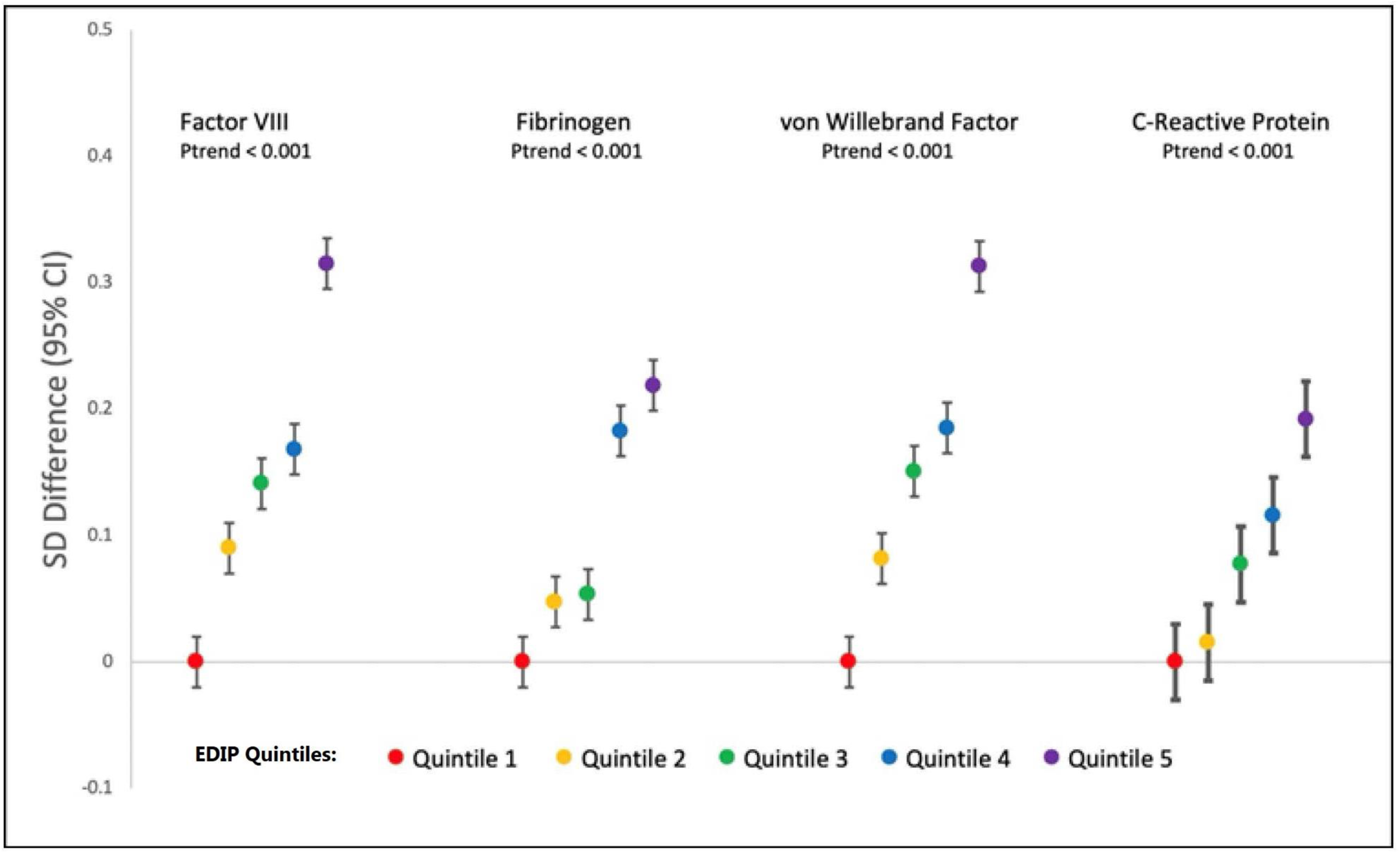
Mean standardized (z-score) levels of factor VIII, fibrinogen, von Willebrand factor (vWF), and C-reactive protein (CRP) are displayed across quantiles of Empirical Dietary Inflammatory Potential (EDIP). Higher EDIP scores (more pro-inflammatory diets) were associated with stepwise increases in all inflammatory biomarkers.

### EDIP and Risk of AF

Over a median 24.2 years of follow up, 1453 participants developed AF (incidence rate 8.6 per 1,000 person-years) with the highest incidence observed among individuals in the highest EDIP quantile **(Figure 2)**. Restricted cubic spline analyses demonstrated a positive dose–response association between EDIP score and incident atrial fibrillation, with no evidence of non-linearity **(Figure 3).** In Cox proportional hazards models (**Table 2**), higher EDIP score was associated with increased risk of incident AF in models adjusted for sociodemographic and traditional cardiovascular risk factors (Model 2 HR 1.21; 95% CI 1.03–1.43). In a continuous fashion, each one–standard deviation increase in EDIP score was associated with higher risk of AF, however, adjustment for cardiovascular risk factors attenuated the association and did not reach statistical significance. Of note, those with incident AF had higher levels of all inflammatory biomarkers at baseline **(Supplementary Table 1)**.

**Figure 2:**
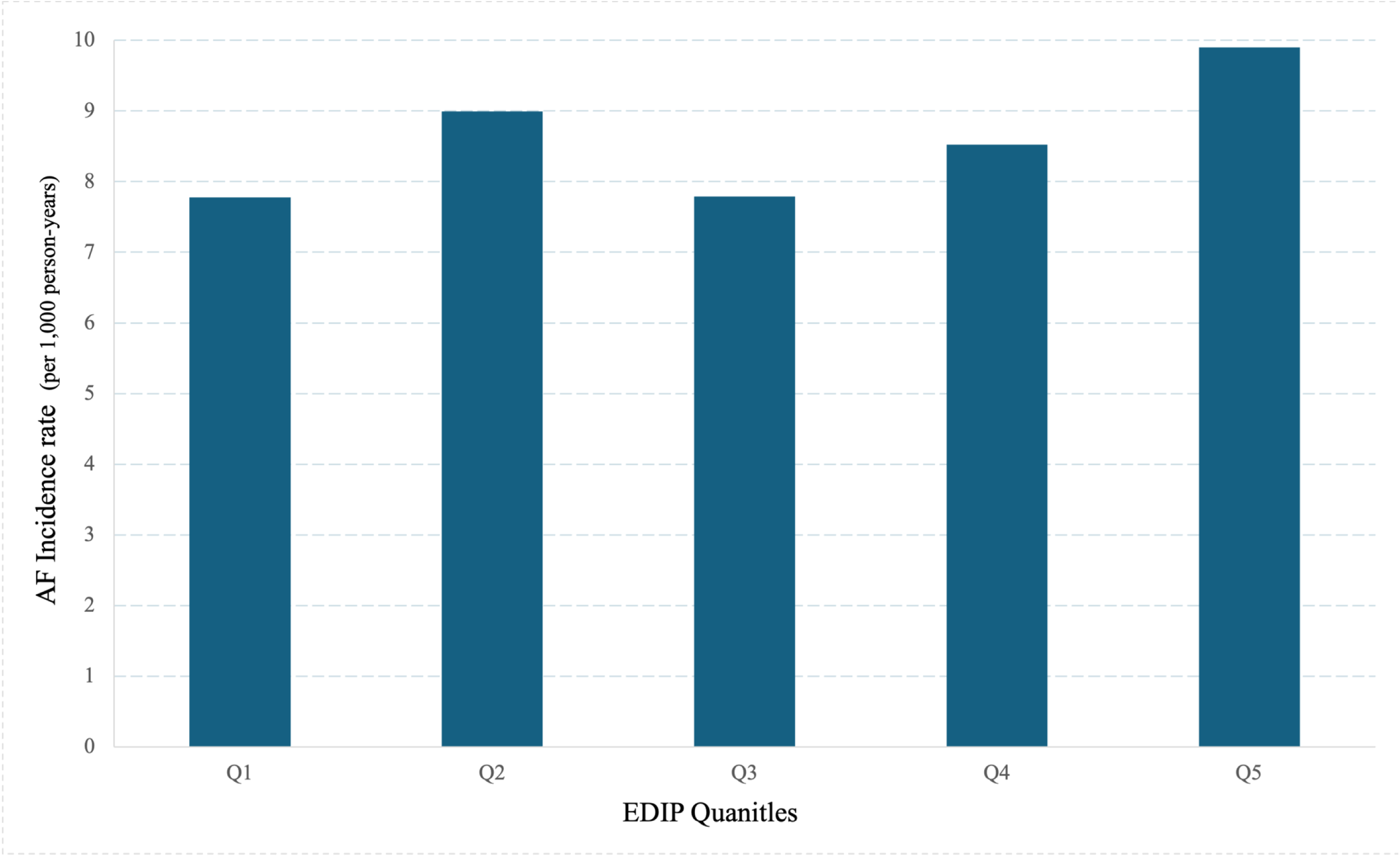
Atrial fibrillation incidence rates (per 1,000 person-years) across EDIP quantiles are shown. Participants with the most pro-inflammatory diets (highest EDIP quantile) experienced the highest incidence of atrial fibrillation during follow-up

**Figure 3:**
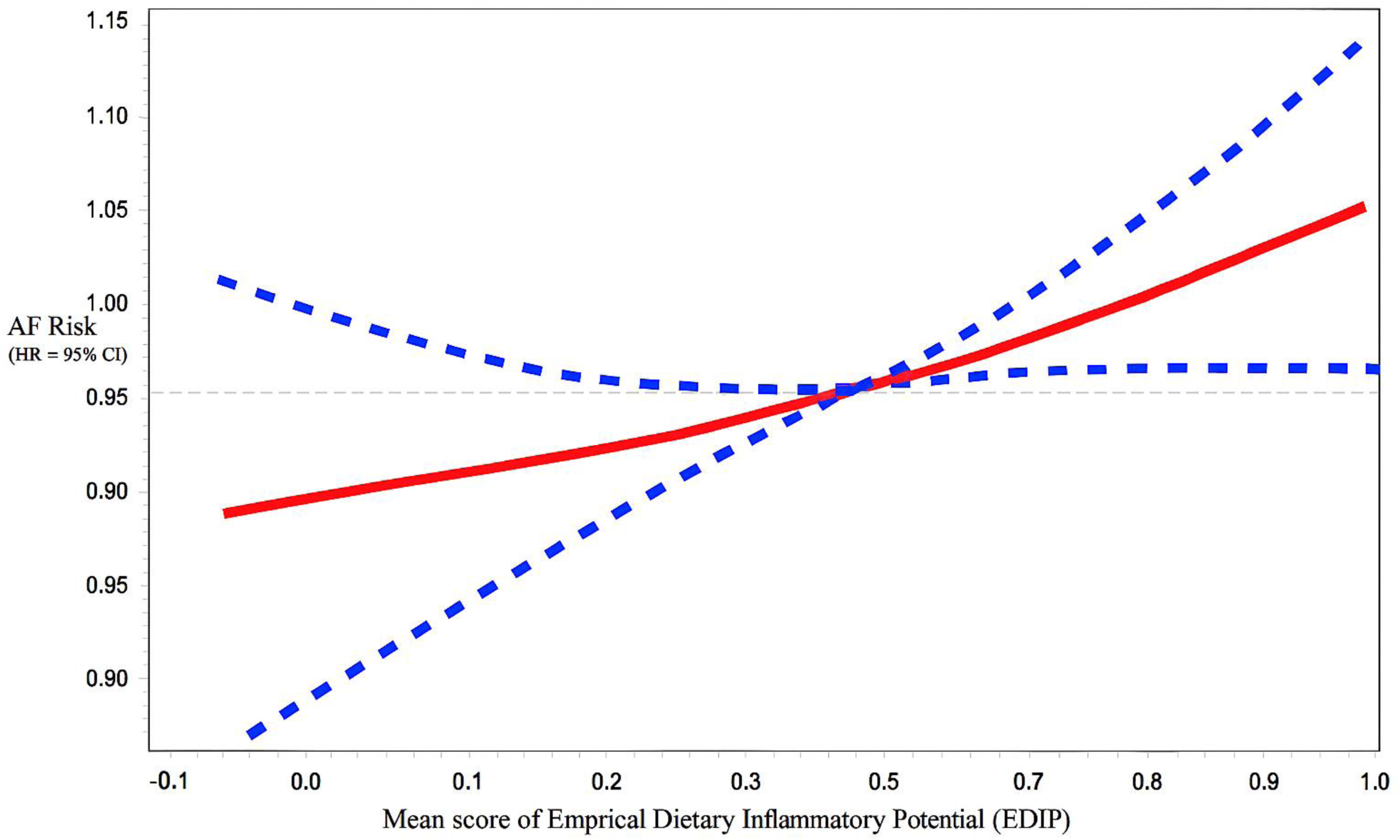
Restricted cubic spline plot illustrating the adjusted hazard ratio for incident atrial fibrillation (AF) across the continuous Empirical Dietary Inflammatory Potential (EDIP) score distribution. The solid red line represents the estimated hazard ratio, and the blue dashed lines indicate the 95% confidence interval. A positive linear association was observed, with higher EDIP scores associated with higher AF risk.

**Table 2:**
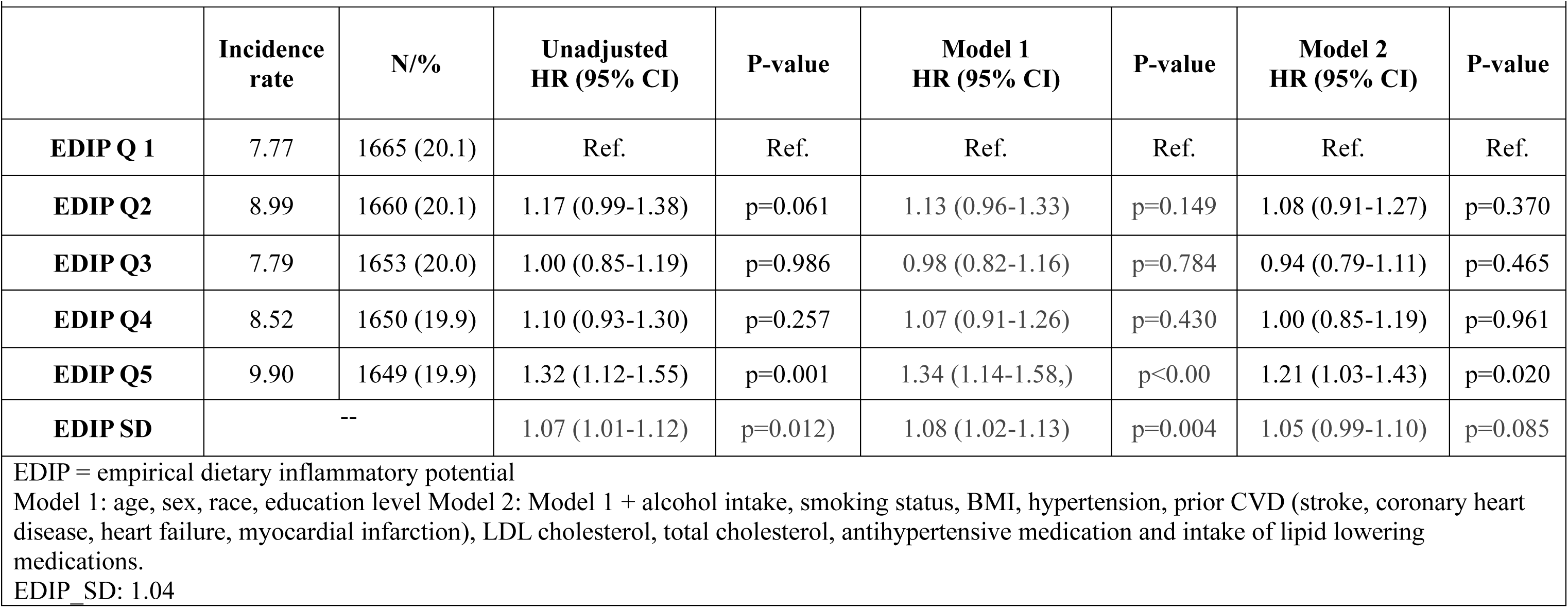
EDIP Score and Risk of AF.

In sex-stratified analyses, the association between EDIP and AF differed between men and women **(Tables 3 and 4**) was examined. Among women, neither EDIP quantiles nor EDIP as a continuous variable were significantly associated with incident AF. In contrast, among men, higher EDIP was consistently associated with greater AF risk. In adjusted models, men in the highest EDIP quantile had a 43% higher risk of AF compared with those in the lowest quantile (Model 2, HR 1.43; 95% CI 1.14–1.79). Each one–standard deviation increase in EDIP was also associated with a 9% increased AF risk among men.

**Table 3:** EDIP and Risk of AF in Women.

|  | **N. of AF**<br>**case**<br>**582** | **Unadjusted/univariate**<br>**HR (95% CI)** | **P-**<br>**value** | **Model 1**<br>**HR (95% CI)** | **P-**<br>**value** | **Model 2**<br>**HR (95% CI)** | **P-**<br>**value** |
| --- | --- | --- | --- | --- | --- | --- | --- |
| **EDIP Q 1** | 114 | Ref. | Ref. | Ref. | Ref. | Ref. | Ref. |
| **EDIP Q2** | 122 | 1.11 (0.86–1.43) | 0.436 | 1.06 (0.82–1.36) | 0.68 | 1.00 (0.78-1.29) | 0.988 |
| **EDIP Q3** | 118 | 1.02 (0.79–1.32) | 0.882 | 0.99 (0.76–1.28) | 0.914 | 0.97 (0.75-1.25) | 0.787 |
| **EDIP Q4** | 123 | 1.10 (0.86–1.42) | 0.449 | 1.10 (0.85–1.42) | 0.47 | 1.01 (0.78-1.30) | 0.966 |
| **EDIP Q5** | 105 | 1.05 (0.81–1.37) | 0.697 | 1.11 (0.85–1.45) | 0.45 | 1.02 (0.78-1.33) | 0.879 |
| **EDIP SD** | -- | 1.01 (0.92–1.09) | 0.651 | 1.02 (0.94–1.12) | 0.289 | 1.01 (0.92–1.10) | 0.85 |

HR 95%CI are derived from Cox proportional hazards models with time-to-incident AF as the outcomes among Women only
EDIP = empirical dietary inflammatory potential.
EDIP\_SD: 1.04
Model 1: age, race, education Model 2: Model 1 + alcohol intake, smoking status, BMI, hypertension, prior CVD (stroke, coronary heart disease, heart failure, myocardial infarction), LDL cholesterol, total cholesterol, antihypertensive medication and intake of lipid lowering medications.

**Table 4:** EDIP and Risk of AF in Men.

|  | **N. of AF**<br>**case**<br>**871** | **Unadjusted/uni**<br>**variate**<br>**HR (95% CI)** | **P-value** | **Model 1**<br>**HR (95% CI)** | **P-value** | **Model 2**<br>**HR (95% CI)** | **P-value** |
| --- | --- | --- | --- | --- | --- | --- | --- |
| **EDIP Q1** | 155 | Ref. | Ref. | Ref. | Ref. | Ref. | Ref. |
| **EDIP Q2** | 185 | 1.21 (0.98–1.50) | 0.081 | 1.18 (0.96–1.47) | 0.123 | 1.20 (0.96–1.51) | 0.116 |
| **EDIP Q3** | 149 | 1.00 (0.80–1.25) | 0.989 | 0.97 (0.77–1.21) | 0.777 | 0.92 (0.72–1.17) | 0.488 |
| **EDIP Q4** | 166 | 1.10 (0.89–1.37) | 0.384 | 1.06 (0.85–1.31) | 0.629 | 1.07 (0.85–1.36) | 0.564 |
| **EDIP Q5** | 216 | 1.47 (1.19–1.80) | 0.0003 | 1.50 (1.22–1.84) | 0.0001 | 1.43 (1.14–1.79) | 0.0018 |
| **EDIP SD** | -- | 1.091 (1.026-1.160) | 0.005 | 1.108 (1.040-1.180) | 0.0015 | 1.092 (1.020-1.170) | 0.012 |

HR 95%CI are derived from Cox proportional hazards models with time-to-incident AF as the outcomes among Men only
EDIP = empirical dietary inflammatory potential.
EDIP\_SD: 1.04
Model 1: age, race, education Model 2: Model 1 + alcohol intake, smoking status, BMI, hypertension, prior CVD (stroke, coronary heart disease, heart failure, myocardial infarction), LDL cholesterol, total cholesterol, antihypertensive medication and intake of lipid lowering medications.

Stratified analysis was performed based on subgroups and HR with 95% CI per 1 SD increase was obtained (**Table 5**). The association between EDIP (per 1 SD increase) and AF risk was overall consistent across subgroups of race, sex, age, diabetes status, and smoking status (interaction p-values > 0.05). A statistically significant interaction was observed for BMI category (interaction p = 0.015): the association of EDIP and risk of AF was more pronounced among obese participants, whereas an inverse association was observed among participants with normal BMI.

**Table 5:** Stratified Analysis.

| Table 5: Stratified Analysis |  |  |  |
| --- | --- | --- | --- |
| Sub-group |  | Hazard Ratio<br>(95% Confidence Interval) * | Interaction *p*-value<sup>‡</sup> |
| Race | *Blacks* | 1.17 (1.02 – 1.35) | 0.081 |
|  | *White* | 1.02 (0.97 – 1.09) |  |
| Gender | *Men* | 1.07 (1.00 – 1.14) | 0.2047 |
|  | *Women* | 1.00 (0.91 – 1.09) |  |
| Age | < 65 | 1.04 (0.99 – 1.10) | 0.74 |
|  | ≥ 65 | 1.17 (0.60 – 2.32) |  |
| BMI classification | Normal (BMI 18.5–24.9) | 0.46 (0.24 – 0.88) | 0.015 |
|  | Overweight (BMI 25.0–29.9) | 1.01(0.93 – 1.09) |  |
|  | Obese (BMI ≥30) | 1.14 (1.04 – 1.25) |  |
| Diabetes | *Present* | 1.09 (0.93 – 1.28) | 0.52 |
|  | *Absent* | 1.03 (0.97 – 1.09) |  |
| Smoking Status | *Current* | 1.09 (0.998 – 1.18) | 0.46 |
|  | *Former* | 1.01 (0.88 – 1.15) |  |
|  | *Never* | 1.07 (0.87 – 1.12) |  |
| Model are adjusted for age, race, gender, education, Model 2: Model 1 + alcohol intake, smoking status, BMI, hypertension, prior CVD (stroke, coronary heart disease, heart failure, myocardial infarction) , LDL cholesterol, total cholesterol, antihypertensive medication and intake of lipid lowering medications.<br>*Hazard ratios are per 1 SD increase in EDIP score.<br>EDIP SD: 1.04 |  |  |  |

## DISCUSSION

In this analysis in the ARIC Study, a middle-aged biracial general cohort, pro-inflammatory dietary patterns, as reflected by higher EDIP scores, was correlated with inflammatory markers at baseline, and associated with increased risk of incident AF after adjustment for sociodemographic and CV risk factors over a median 24.2 years of follow-up. Furthermore, sex-stratified analysis revealed a significant sex-difference: a significant association observed among men but not among women. Obese participants had more pronounced effect of EDIP on the risk of AF.

### Diet, Inflammation and Risk of AF

Previous epidemiologic studies have consistently shown that elevated inflammatory biomarkers, particularly CRP, predict both prevalent and incident AF in community cohorts, suggesting that low-grade systemic inflammation may precede arrhythmia onset.^14,17,33^ An analysis in the Framingham Heart Study extended these findings by showing that inflammatory markers including CRP, interleukin-6 (IL-6), fibrinogen, and tumor necrosis factor receptors were jointly associated with incident AF, indicating that a broader inflammatory state underlies AF susceptibility.^34^

Chronic exposure to these inflammatory mediators promotes AF through structural, conductive, and electrical remodeling of the atria through cytokine-driven fibrosis, extracellular matrix deposition, myocyte apoptosis, disruption of gap junction integrity, and alterations in ion channel function creating a substrate for reentry and ectopic activity.^35–38^ For these inflammatory markers to exert effects on cardiac tissue and promote arrhythmogenesis, chronic exposure is likely required to produce clinically significant disease. Therefore, identifying upstream pathways that influence systemic inflammation may identify mechanistic links and opportunities for targeted preventive strategies.

Dietary habits represent a key upstream determinant of systemic inflammation. Studies in both the UK Biobank and REGARDS reported lower AF risk among individuals adhering to heart-healthy patterns such as the Mediterranean and DASH diets; however, these associations frequently attenuate after adjustment for adiposity and other lifestyle factors. ^39,40^ Consistent with these observations, recent systematic reviews showed that evidence supporting specific dietary patterns for primary AF prevention remains inconclusive except for alcohol intake.^41^

One plausible explanation for this inconsistency in predictability of traditional diet-quality indices, at least in part, is the differences in how dietary exposure operationalized and that these indices primarily capture cardiometabolic benefits while overlooking the inflammatory potential of dietary intake.^42^ The EDIP score represents a mechanistically informed approach that was specifically developed to quantify the inflammatory influence of diet.^43^ In our study, higher EDIP scores showed clear positive correlations with all measured inflammatory biomarkers at baseline and demonstrated dose–response relationships with incident AF underscoring the potential value of dietary inflammatory potential, beyond traditional diet-quality metrics alone, in understanding and predicting AF risk.

### Potential Pathophysiological Mechanisms

Our observations suggest that dietary patterns influence AF risk through chronic inflammatory pathways. This is evident by the positive dose–response relationship observed in spline analyses and the incremental increase in AF risk with higher EDIP quantiles. While generalized diet-quality indices such as the Mediterranean Diet Score and DASH diet score may be useful for assessing cardiometabolic health, EDIP, which explicitly capture dietary inflammatory load, might offer a supplementary biological relevance for AF pathogenesis and prevention.^41,44^

Supporting this framework, interventional and secondary prevention studies have suggested benefits of anti-inflammatory dietary approaches, including Mediterranean diets, particularly when enriched with extra-virgin olive oil, in both general populations and post-AF ablation settings.^45,46^ Collectively, our results position EDIP as a distinct and modifiable upstream factor in AF pathogenesis and could provide an actionable pathway to reduce AF risk through targeted dietary strategies that lower chronic inflammatory exposure.

### Sex Difference between EDIP and risk of AF

Our study revealed a significant association between dietary inflammatory potential and AF risk among men, but not women. This aligns with known sex differences in AF epidemiology, where men develop AF earlier and more frequently, while women present later and with more advanced disease.^47,48^ Emerging evidence suggests that upstream AF risk factors may operate differently by sex, particularly with respect to inflammation and modifiable lifestyle exposures.^10,48–50^ In the VITAL Rhythm Study, cardiometabolic and inflammatory predictors showed larger relative associations with AF in men, and biomarker data from the ISOLATION AF registry demonstrated that men exhibit stronger inflammatory and thrombogenic profiles, whereas women show greater fibrosis and endothelial dysfunction.^51,52^

These findings suggest that dietary inflammatory load, as captured by EDIP, may contribute more prominently to AF risk in men through systemic inflammatory pathways.^14^ In contrast, AF in women appears to manifest later in life and in the setting of more advanced structural remodeling, potentially diminishing the relative contribution of upstream dietary inflammatory exposures^.53^ These observations underscore the importance of considering sex-specific mechanisms when evaluating diet–inflammation pathways and support dietary inflammation reduction as a potentially more relevant preventive strategy for men.

### Obesity as an Effect Modifier

We observed a significant interaction between BMI and EDIP, with the strongest associations between dietary inflammatory potential and AF among individuals with obesity (BMI ≥30), whereas the relationship was attenuated or even inverse among normal-weight participants. Given that obesity is a major AF risk factor, these results highlight a synergistic effect in which pro-inflammatory diets may compound obesity-related atrial susceptibility. ^54,55^

Obesity is characterized by chronic low-grade inflammation and secretion of pro-inflammatory adipokines, which might act synergistically with pro-inflammatory dietary patterns to accelerate atrial remodeling through mechanisms such as atrial enlargement and fibrosis.^56^ Importantly, EDIP correlates positively with BMI and other adiposity measures, indicating that pro-inflammatory diets often coexist with excess body weight. ^57^ This interplay underscores the importance of weight management as a cornerstone of AF prevention and suggests that dietary strategies reducing inflammatory load would confer additional benefit in obese individuals.

### Clinical and Public Health Implications

Our findings underscore the importance of dietary patterns as a modifiable risk factor for AF. The observed association between EDIP and AF, particularly among men and individuals with obesity, suggests that reducing dietary inflammatory load could complement existing prevention strategies focused on weight management and cardiovascular risk control.

Incorporating dietary assessment into AF risk stratification may enable more personalized interventions. Public health initiatives promoting anti-inflammatory dietary patterns such as increased intake of fruits, vegetables, and whole grains could have an impact on AF prevention at the population level.

### Strengths and Limitations

The strengths of our study include its large, community-based cohort, long-term follow-up, rigorous AF ascertainment, comprehensive adjustment for confounders and subgroup analyses allowing for exploration of effect modification. However, several limitations should be acknowledged. First, observational design precludes causal inference, and residual confounding cannot be excluded. Second, dietary intake was measured only at baseline, limiting our ability to capture changes over time. Finally, misclassification of dietary exposures was possible. Despite using the original coefficients from Tabung et al.^19^ without modification to calculate the EDIP, minor differences in food-group categorization might have existed to align with ARIC FFQ structure. ^24^

### Conclusions

In conclusion, pro-inflammatory dietary patterns were associated with increased risk of AF in a middle-aged biracial general cohort. These findings provide insights into the interplay between diet-related inflammation and AF risk, highlighting potential pathophysiological pathways and targeted preventive strategies through dietary modification of inflammatory pathways.

## Data Availability

The data used in this study were obtained from the National Heart, Lung, and Blood Institute (NHLBI) Biologic Specimen and Data Repository Information Coordinating Center (BioLINCC). The datasets used for this analysis are available under the ARIC BioLINCC accession number HLB00020025a. Researchers may request access to these data directly through the BioLINCC website.

## Acknowledgments

The Atherosclerosis Risk in Communities Study is carried out as a collaborative study supported by National Heart, Lung, and Blood Institute (NHLBI) contracts (HHSN268201700001I, HHSN268201700002I, HHSN268201700003I, HHSN268201700004I, and HHSN268201700005I). The authors thank the staff and participants of the ARIC study for their important contributions. This manuscript was prepared using a limited access dataset obtained from the NHLBI Biologic Specimen and Data Repository Information Coordinating Center (BioLINCC) and does not necessarily reflect the opinions or views of the ARIC study investigators or the NHLBI.

## ABBREVIATIONS

AF: atrial fibrillation
ARIC: Atherosclerosis Risk In Communities
BMI: body mass index
ECG: electrocardiogram
EDIP: empirical dietary inflammatory potential
FFQ: food frequency questionnaire

## Notes

### Competing Interest Statement

The authors have declared no competing interest.

### Funding Statement

this research did not receive any external funds. The Atherosclerosis Risk in Communities Study is carried out as a collaborative study supported by National Heart, Lung, and Blood Institute (NHLBI) contracts (HHSN268201700001I, HHSN268201700002I, HHSN268201700003I, HHSN268201700004I, and HHSN268201700005I). The authors thank the staff and participants of the ARIC study for their important contributions. This manuscript was prepared using a limited access dataset obtained from the NHLBI Biologic Specimen and Data Repository Information Coordinating Center (BioLINCC) and does not necessarily reflect the opinions or views of the ARIC study investigators or the NHLBI.

### Author Declarations

this manuscript used public access dataset. No further IRB approval needed as data was daintified as the time of usage.

